# SARS-CoV-2 infection induces sustained humoral immune responses in convalescent patients following symptomatic COVID-19

**DOI:** 10.1101/2020.07.21.20159178

**Authors:** Jun Wu, Boyun Liang, Cunrong Chen, Hua Wang, Yaohui Fang, Shu Shen, Xiaoli Yang, Baoju Wang, Liangkai Chen, Qi Chen, Yang Wu, Jia Liu, Xuecheng Yang, Wei Li, Bin Zhu, Wenqing Zhou, Huan Wang, Shumeng Li, Sihong Lu, Di Liu, Huadong Li, Adalbert Krawczyk, Mengji Lu, Dongliang Yang, Fei Deng, Ulf Dittmer, Mirko Trilling, Xin Zheng

**Affiliations:** Department of Infectious Diseases, Union Hospital, Tongji Medical College, Huazhong University of Science and Technology, Wuhan 430022, China; Joint International Laboratory of Infection and Immunity, Huazhong University of Science and Technology, Wuhan 430022, China; Department of ICU, Fujian Medical University Union Hospital, Fuzhou, 350001, China; State Key Laboratory of Virology, Wuhan Institute of Virology, Chinese Academy of Sciences, Wuhan 430071, China; Ministry of Education Key Lab of Environment and Health, School of Public Health, Tongji Medical College, Huazhong University of Science and Technology, Wuhan 430030, China; Hubei Provincial Center for Disease Control and Prevention, Wuhan 430079, China; Pritzker School of Medicine, University of Chicago, Chicago, USA; Jin Yin-tan Hospital, Wuhan 430022, China; Department of Infectious Diseases, University Hospital of Essen, University of Duisburg-Essen, Essen 45147, Germany; Institute for Virology, University Hospital of Essen, University of Duisburg-Essen, Essen 45147, Germany

**Keywords:** COVID-19, SARS-CoV-2, humoral immunity, antibody responses, sustained immunity

## Abstract

Long-term antibody responses and neutralizing activities following SARS-CoV-2 infections have not yet been elucidated. We quantified immunoglobulin M (IgM) and G (IgG) antibodies recognizing the SARS-CoV-2 receptor-binding domain (RBD) of the spike (S) or the nucleocapsid (N) protein, and neutralizing antibodies during a period of six months following COVID-19 disease onset in 349 symptomatic COVID-19 patients, which were among the first world-wide being infected. The positivity rate and magnitude of IgM-S and IgG-N responses increased rapidly. High levels of IgM-S/N and IgG-S/N at 2-3 weeks after disease onset were associated with virus control and IgG-S titers correlated closely with the capacity to neutralize SARS-CoV-2. While specific IgM-S/N became undetectable 12 weeks after disease onset in most patients, IgG-S/N titers showed an intermediate contraction phase, but stabilized at relatively high levels over the six months observation period. At late time points the positivity rates for binding and neutralizing SARS-CoV-2-specific antibodies was still over 70%. Taken together, our data indicate sustained humoral immunity in recovered patients who suffer from symptomatic COVID-19, suggesting prolonged immunity.

## Introduction

As of July 20, 2020, the global number of confirmed cases of Corona Virus Disease 2019 (COVID-19) has reached 14,3 million, with more than 603,691 known fatalities. In December 2019, the sarbecovirus *severe acute respiratory syndrome coronavirus 2* (SARS-CoV-2) was identified as causative pathogen causing COVID-19 ^1^. The virus has spread around the world at a rapid pace. The COVID-19 pandemic represents the greatest medical and socio-economic challenge of our time. There is neither a sufficiently effective antiviral drug to treat COVID-19 cases nor an approved vaccine. It is crucial for decision-making and vaccine development to understand how long immunity against SARS-CoV-2 persists in infected individuals and whether antibodies produced in response to a natural infection provide protective immunity, which may prevent re-infection with SARS-CoV-2.

To our knowledge, the longest observation period for SARS-CoV-2 specific antibodies has only been 12 weeks ^2^ and it remains unclear how antibody titers may change over subsequent periods. Due to the use of different detection methods (e.g., ELISA versus CLIA), the analysis of different subtypes of antibodies (IgG, IgM or IgA) and the focus on different antigens and epitopes (N, S or the receptor binding domain [RBD] of S), a coherent description of the humoral immune response after natural SARS-CoV-2 infections is not available. As has been consistently shown in short-term studies, a seroconversion of IgG and IgM occurs about two to three weeks after disease onset^3^ and IgM levels drop significantly earlier than IgG titers ^4^. However, it is unclear which antibody type (IgG or IgM) performs best in the epidemiologic identification of convalescent patients. Some authors favored IgG ^3,4^, while other proposed a higher positivity rate for IgM ^5^. In addition, the reported peak of IgM responses was assigned to different time points ranging from two to five weeks ^2,3,5^. Thus, we set out to clarify the kinetics and magnitude of the initial antibody response against SARS-CoV-2 in a large cohort of symptomatic COVID-19 patients from Wuhan.

Most importantly, many of these patients, which were among the first becoming infected with SARS-CoV-2 world-wide, were followed up for several months to determine how sustainable the antibody response against SARS-CoV-2 is. So far studies that analyzed only a few patients or that had an observation period of only a few weeks, suggested that antibody levels may decrease rapidly in infected individuals ^6^. This has been greatly discussed world-wide because it may be a very important aspect for natural immunity and vaccine development. However, long-term studies are needed because immune responses always decline after acute infections which does not predict the duration of a protective response.

SARS-CoV-2 has a single stranded positive-sense RNA genome which encodes structural and nonstructural proteins, including the spike (S) and the nucleocapsid (N) protein ^7^. A part of the transmembrane S protein is present on the virion surface and binds to the entry receptor ACE2 mediating entry into target cells ^8^, while the highly abundant N protein binds to the viral RNA inside viral particles. Previous research on SARS and MERS has shown that IgG responses recognizing S and N have different characteristics in terms of response time, duration, and titers ^9,10^. In certain diseases such as Dengue virus infections, binding but non-neutralizing antibodies have even been associated with worse clinical outcomes through antibody-dependent enhancement (ADE), suggesting that under certain circumstances antibodies may at least correlate with harmful effects in some patients. ADE in the context of SARS-CoV-2 has been discussed recently ^11^. Higher antibodies have also been associated with older age ^2^ in COVID-19 patients. However, studies using pseudovirus particle-based systems ^12,13^,suggest that plasma derived from convalescent patients have potent neutralizing activity that was related to IgG molecules recognizing the RBD of the S protein, suggesting that IgG-RBD-S antibodies have a high likelihood to fulfill neutralizing functions (nAbs). Some small cohort studies suggest that severe COVID-19 patients benefit from convalescent plasma (CP) therapy ^14^. Very recently, highly potent SARS-CoV-2 neutralizing antibodies have been isolated and characterized from COVID-19 patients^15,16^. Thus, virus-specific antibodies seem to be very important for immunity against SARS-CoV-2 infection and/or COVID-19 development. However, it remains to be clarified how the kinetics of binding and neutralization antibodies change during and after the course of COVID-19.

The aim of this study was to evaluate the IgM and IgG responses against the RBD of the S protein and the nucleocapsid protein (N) longitudinally after the onset of symptomatic COVID-19. Presence of these antibodies and neutralizing activities of plasma were determined throughout a period of 26 weeks. The results of the study shall provide an experimental basis for evaluating the onset and duration of humoral immunity in COVID-19 patients in order to support clinical drug and vaccine development and decision making in terms of social-economic mitigation strategies.

## Results

### Symptomatic COVID-19 patients exhibit an early and rapid IgM-S and IgG-N response and maintain high levels of IgG-S/N for at least six months after disease onset

In order to investigate antibody responses towards SARS-CoV-2 over time, a total of 585 samples obtained from 349 symptomatic COVID-19 patients, collected up to 26 weeks after disease onset, were analyzed for IgM and IgG recognizing the RBD of the spike protein (denoted IgM-S and IgG-S, respectively) as well as IgM and IgG binding the nucleocapsid protein (IgM-N and IgG-N, respectively). As test system a capture chemiluminescence immunoassays was used.

During the initial outbreak in Wuhan, nucleic acid based detection methods were always complemented with antibody detection assays for the diagnosis of suspected COVID-19 patients. All analyzed patients in this study were symptomatic for COVID-19. During the first week after symptom onset, the four antibodies were tested positive with different frequencies: IgM-S (66%) > IgG-N (33%) > IgM-N (22%) > IgG-S (11%) (Fig. 1A). The positive rate for IgM-S reached a peak of 94% at week 5 and then rapidly decreased to 0% at week 13 fluctuating below 35% thereafter. IgM-N could be detected in 72% of the patients at week 3. Afterwards, this number rapidly declined and IgM-N became undetectable at week 10 and 12, followed by negligible fluctuations at very low positive rates. IgG-S was already positive in 97% of the patients at week 3 and remained at a relative high percentage until the end of the observation period at week 26. The positive rate of IgG-N rose rapidly to 87% of the patients at week 2 and stayed at very high levels thereafter.

**Fig 1.**
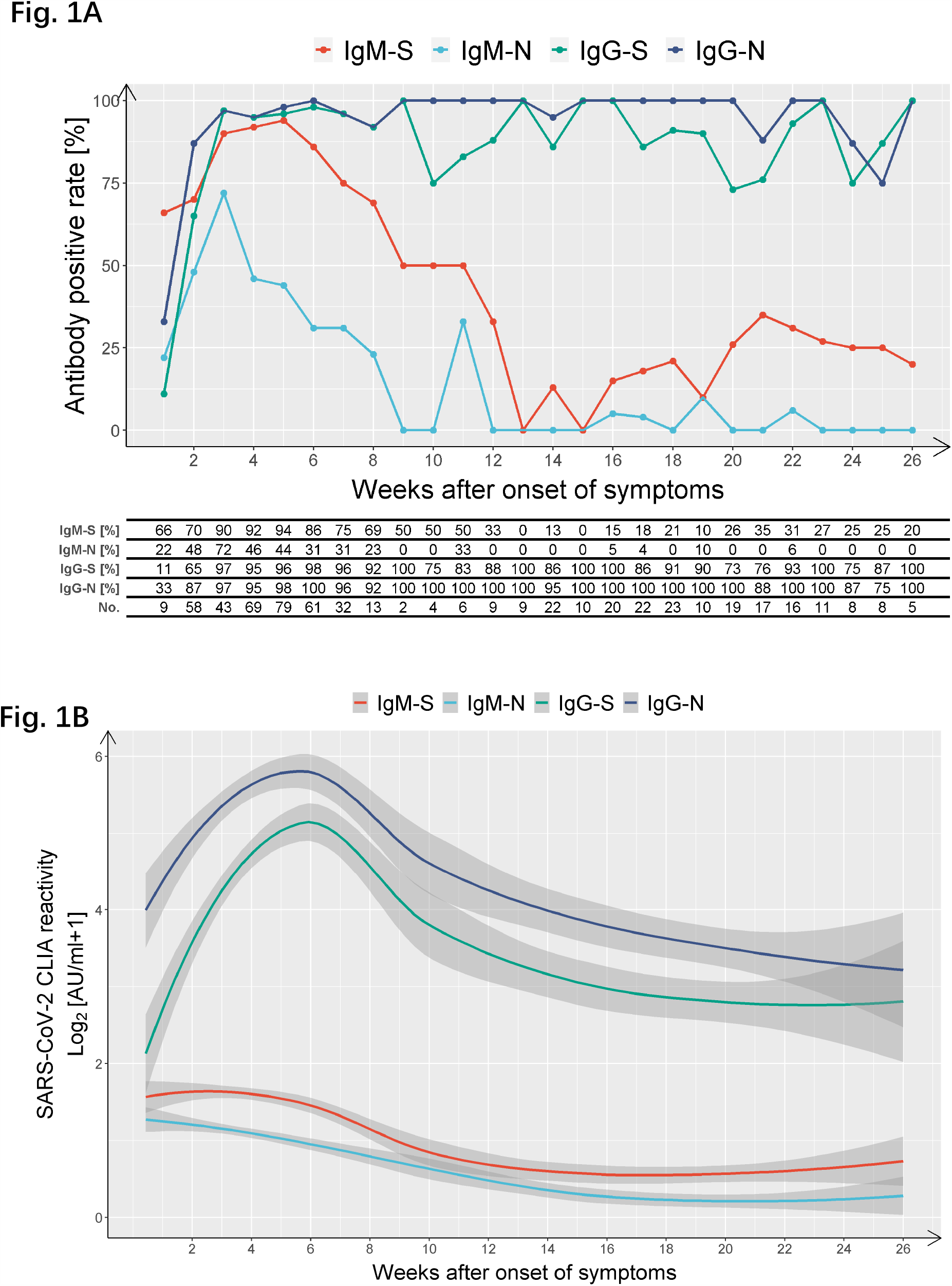
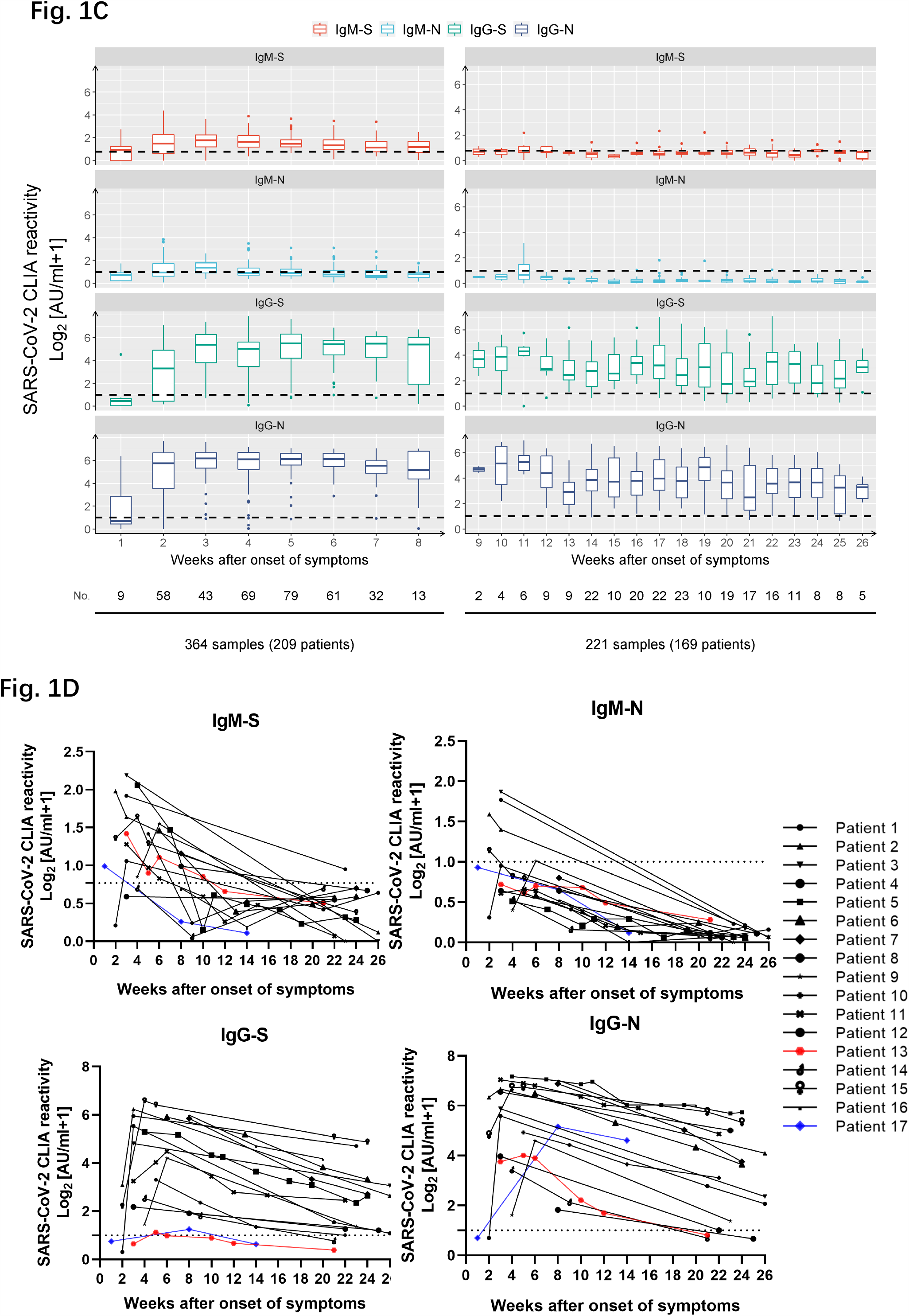
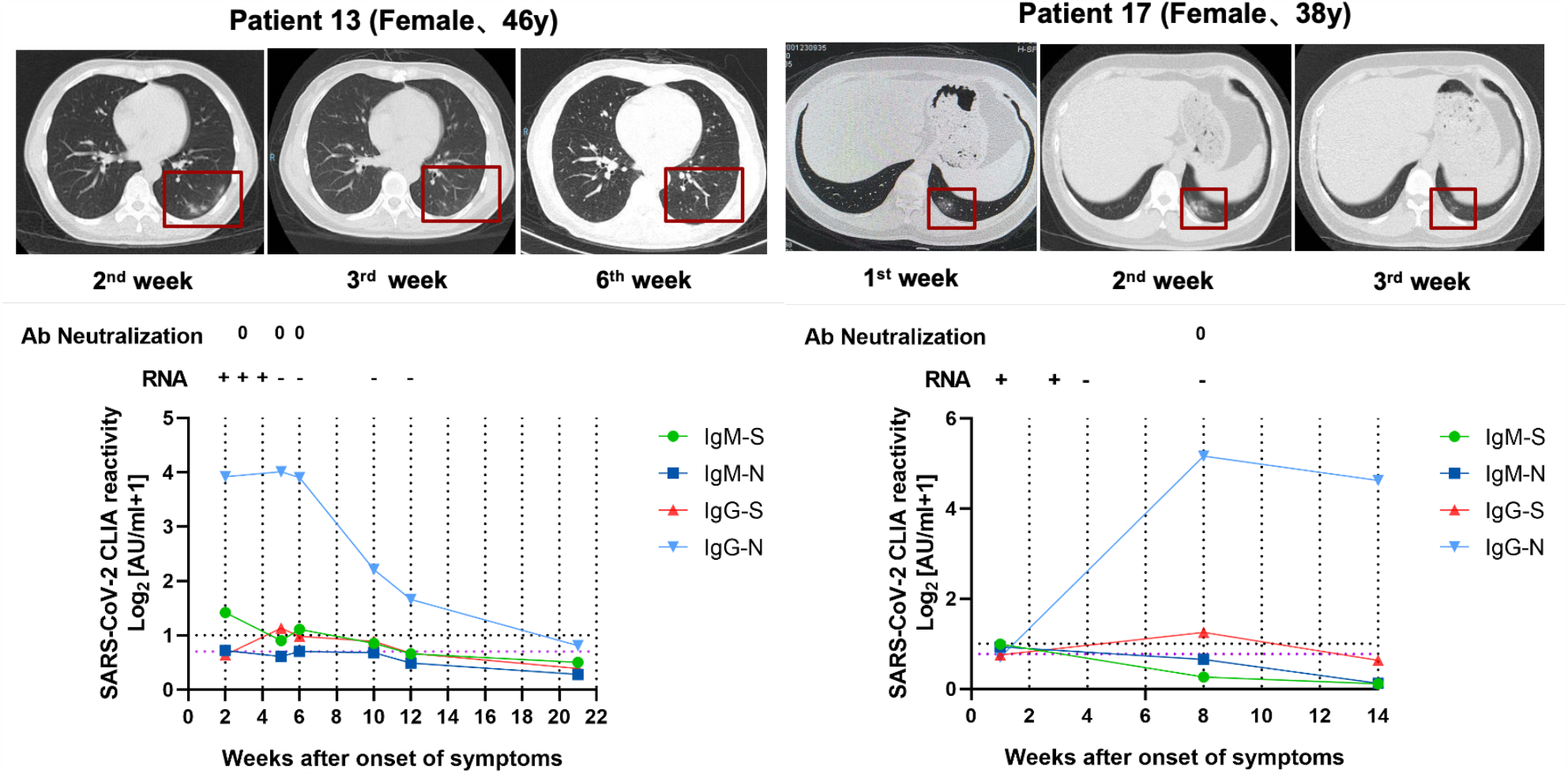
Longitudinal analyses of IgM and IgG responses recognizing the RBD of spike and nucleoproteins of SARS-CoV-2 in confirmed COVID-19 patients. IgM and IgG against the RBD of the spike protein (‘S’) and the nucleoprotein (‘N’) of SARS-CoV-2 were detected by capture chemiluminescence immunoassays (CLIA). (A) Positive rate of individual antibodies tested at the indicated dates following onset of symptoms. (B-C) The plasma antibody levels (IgM-S, IgM-N, IgG-S, and IgG-N) in patients with different disease courses are presented. (D) Sequential sampling and analyses of antibody titers in 17 COVID-19 cases. Characteristics of two patients with low IgG antibody levels. Patient 13: a 46-year-old female with fever, cough, dizziness, and fatigue for 6 days; patient 17: a 38-year-old female with fever and chest tightness for 4 days. The cut-off value for IgM-S detection was 0.7 AU/ml. The cut-off value for IgM-N, IgG-S, and IgG-N were 1 (shown on the left Y axis).

We further analyzed whether a combined antibody tests may support clinical diagnostics (Fig. S1). A combination of IgM-S and IgM-N test did not increase the sensitivity compared to IgM-S alone. Combined IgG-S and IgG-N increased the positive rate compared to IgG-S or IgG-N alone at some time points, suggesting a diagnostic benefit. In agreement with previous studies ^3^, the combination of IgM-S, IgM-N, IgG-S, and IgG-N resulted in positive rates approaching 100% after week 4, indicating that virtually all COVID-19 patients raise detectable humoral immune responses against SARS-CoV-2.

We also determined the dynamics of specific antibody titers during 26 weeks after symptom onset in COVID-19 patients (Fig. 1B and C). Interestingly, IgM-S and IgG-S peaked one week later than IgM-N and IgG-N (Fig. 1B). The titer of IgM-S reached its peak at week 4, and then slowly decreased until the average value fell below the cutoff value at week 12. After reaching the peak at week 3, the titers of IgM-N dropped rapidly below the cutoff value after around 9 weeks. The titers of IgG-N and IgG-S reached their peaks at week 4 and 5, respectively. After a contraction phase, in which titers constantly decreased during week 6 to 14, IgG-N and IgG-S titers stabilized and were maintained at high levels until the end of the observation period of 26 weeks post symptom onset. Thus, SARS-CoV-2-specific IgG responses were very similar to antibody responses against many other viruses with a peak activity a few weeks after infection, which was followed by a contraction phase over several weeks, but finally resulting in a stabilized antibody response that could be detected for at least 6 months.

To corroborate our findings, antibody titers of 17 prototypical patients with repetitive sampling were analyzed. Except for two unusual patients who did not develop a measurable IgG-S response, the same trends in terms of a rapid IgM titer decline and a sustained IgG response after an intermediate contraction were observed (Fig. 1D, upper panel). Intriguingly, the two unusual patients were young women diagnosed with symptomatic COVID-19 accompanied with lung lesions (Fig. 1D, lower panel). Both women exhibited moderate IgG responses recognizing the N protein but did not develop meaningful IgG-S titers. The neutralization activity of their plasma was tested at different time points with consistently negative results, highlighting the importance of S-specific IgG for virus neutralization.

### SARS-CoV-2 is controlled in COVID-19 patients with high levels of IgM-S/N and IgG-S/N at early time points of disease

In order to clarify the interplay between antibodies and virus control, disease severity, gender, as well as age in COVID-19 inpatients, we compared the antibody titers amongst different patient groups. The clinical and laboratory characteristics of COVID-19 patients at the time of admission are depicted in table S1. Taken together, 149 (71.3%) non-severe cases and 60 (28.7%) severe cases from isolation wards with complete medical records were enrolled. No significant differences concerning gender and age were observed between these two groups. Consistent with previous reports^17^, severely ill patients showed significantly decreased counts and frequency of lymphocyte (p<0.01) and decreased PLT counts (p<0.05) compared to patients with non-severe disease courses, while the counts and frequency of neutrophils increased (p<0.01). As expected, patients from the group with severe diseases presented with significantly increased total bilirubin (TBil), alanine aminotransferase (ALT), aspartate transaminase (AST), lactate dehydrogenase (LDH), creatine kinase (CK), Creatinine (Cr), D-dimers, Prothrombin time (PT) and fibrinogen (FIB) than the non-severe group (p<0.05).

In order to investigate the correlation between antibody responses and virus control, patients were stratified according to the presence or absence of SARS-CoV-2 RNA at the time point of antibody determination. At early time points, antibody levels were significantly higher in the group in which SARS-CoV-2 RNA was no longer detected compared to the group with prolonged SARS-CoV-2 RNA positivity. This finding strongly suggests that the presence of IgM and IgG recognizing the S and N protein of SARS-CoV-2 constitutes a clinically relevant correlate of protection in humans and contributes to virus control during the early phase of infection (Fig. 2A & S2A).

**Fig 2:**
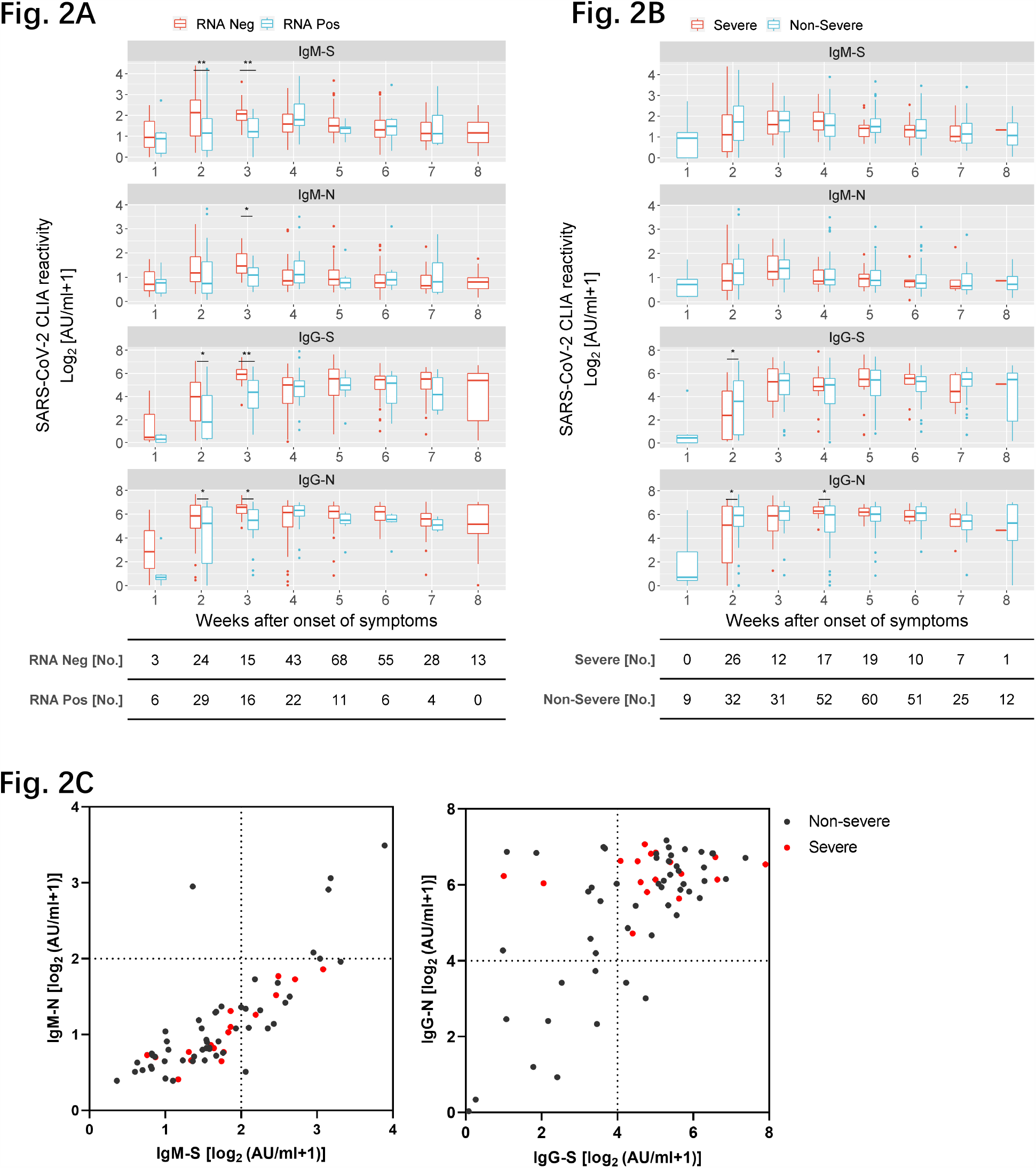
Correlation of antibody titer with virus control and severity of illness in hospitalized COVID-19 patients. (A) S- and N-specific CLIA-reactive IgM/IgG were compared in COVID-19 patients who were virus RNA-negative versus those who were virus RNA-positive at the time-point of sampling at different periods after the disease onset. Each antibody detection value is classified into RNA negative group or RNA positive group according to the simultaneous RNA detection result. A total of 343 results were acquired for this analysis. (B) Comparison of S and N-specific CLIA-reactive IgM/IgG titers between severe and non-severe patients. (C) Comparison of 64 severe and non-severe patients (69 samples) at different S- and N-specific CLIA-reactive IgM/IgG levels at the 4th week after symptoms onset. The whiskers represent the 10th-90th percentiles. GLMM was used for statistical analysis. *p<0.05; **p <0.01; ***p<0.001.

Given the debate concerning the duration of antibody responses in asymptomatic patients ^6^, we wondered if non-severe and severe COVID-19 cases might differ concerning their humoral immune responses. Interestingly, their overall responses were very similar. There were significant higher IgG-S/N responses in patients with non-severe symptoms at weeks 2, again pointing towards a protective role of IgG (Fig. 2B & S2B). The IgG-N levels of patients with severe symptoms were temporarily higher than in those with non-severe disease at week 4 which may be a consequence of higher virus replication and antigen loads raising stronger immune responses. Accordingly, severe patients exhibited high IgG-S and IgG-N titers (Fig.2C).

In general, life-threatening COVID-19 cases are more frequent in males and in the elderly^18^. Therefore, the relationship between gender and age with antibody levels was also investigated. Males tend to have significantly more SARS-CoV-2-specifc IgM (Fig. S2C & S2D), whereas IgG responses did not show a consistent sexual disparity. At later time points, the levels of the four antibodies were significantly higher in elderly patients (≥65 years old) than those in patients younger than 65 years (Fig. S2E & S2F), which might reflect higher viral loads in elderly patients.

### IgG-RBD-S titers correlated closely with the capacity to neutralize SARS-CoV-2

NAbs exhibit strong therapeutic and prophylactic efficacies in SARS-CoV-2-infected hACE2-transgenic mice ^19^ and a recent vaccination study conducted in non-human primates identified Nabs as correlate of protection^20^. In order to study the duration of the neutralization capacity of antibodies, virus neutralization tests were conducted using 186 samples from 137 patients. As early as two weeks post symptom onset, half of the patients demonstrated neutralization activity with at 50% virus neutralization at a minimum plasma dilution of 1:20 (Fig. 3A). By week 4, the proportion of patients with neutralization activity increased to over 90%, and then remained very high until the end of the observation at 26 weeks (Fig. 3A). Neutralizing activity at a serum dilution of 1: 160 has been used as a cutoff in a clinical proof-of-concept study showing the efficacy of CP therapy ^21^. A considerably high frequency of individuals in our study exhibited such strong neutralizing capacities (≥1:160). The finding that elite neutralizers (≥1:320) were not evident before week 7 suggests that it takes some time to raise very potent antibody responses.

**Fig 3:**
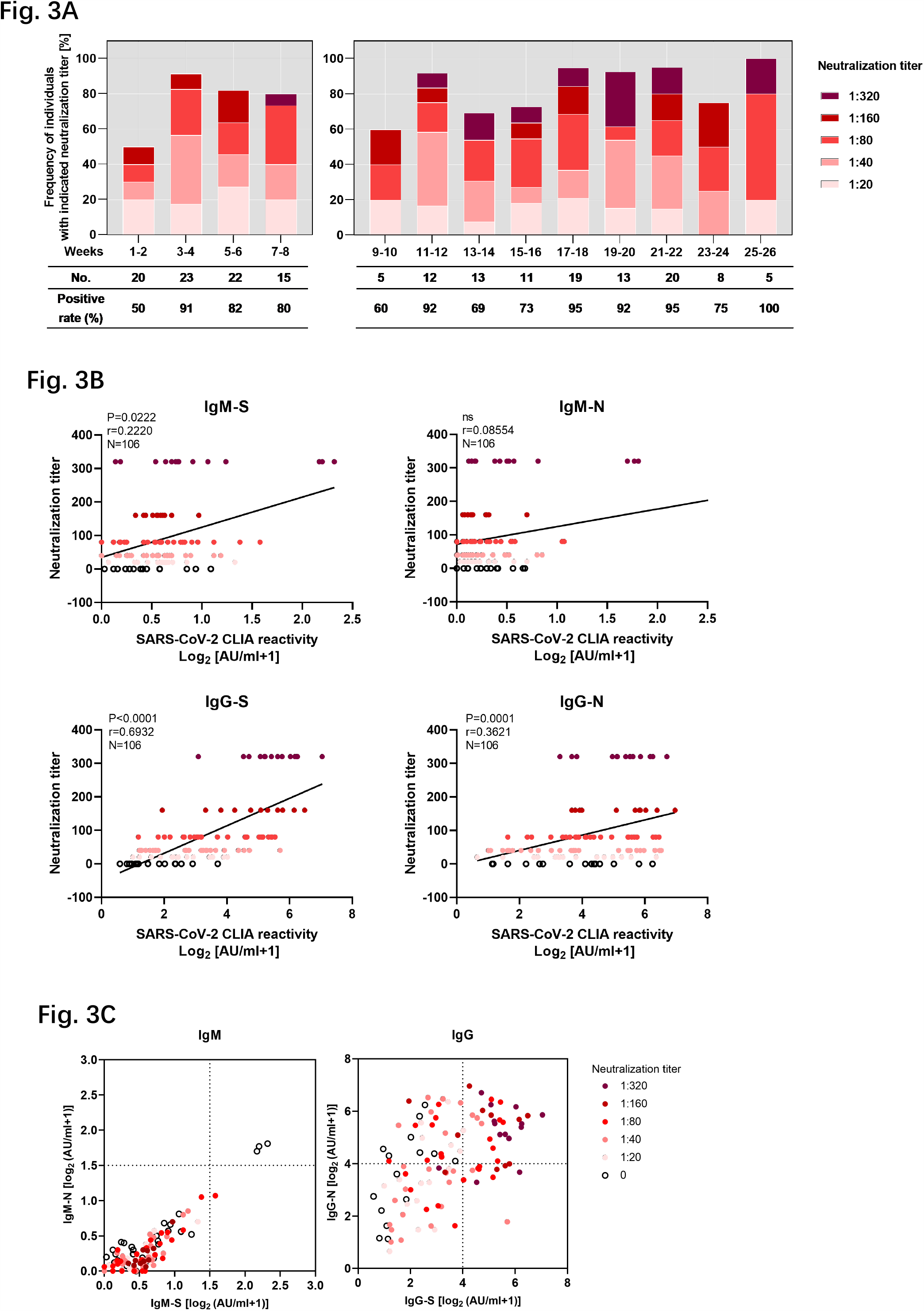
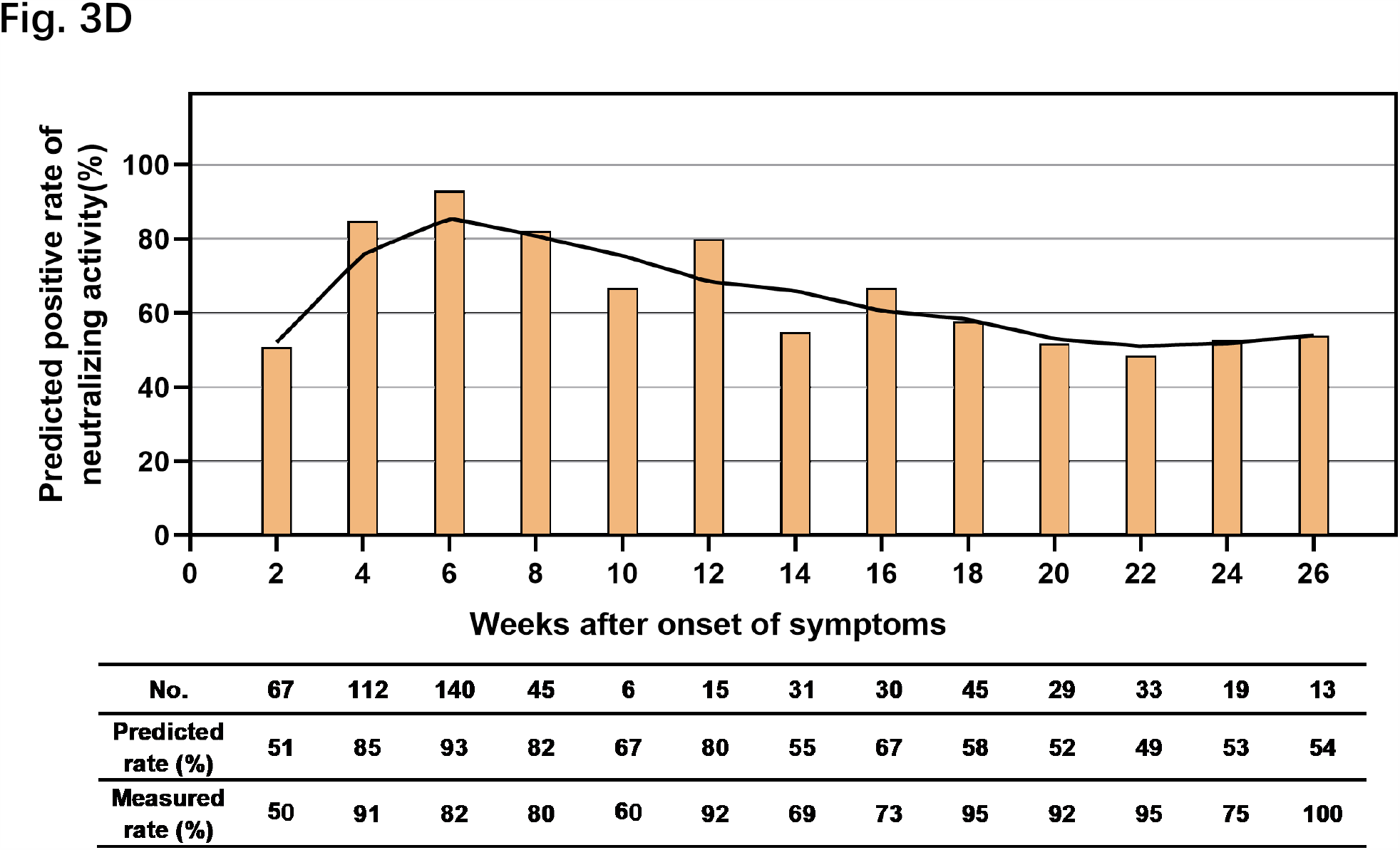
N- and S-specific CLIA-reactive IgG responses have different predictive values for neutralization capacities. A total of 186 samples from 137 symptomatic COVID-19 patients were assessed concerning SARS-CoV-2 neutralization titers and grouped according to the weeks after symptom onset. (A) Proportions of plasma neutralization activity were stratified in tow-week intervals. (B) Correlation analysis of neutralization titer with S- and N-specific CLIA-reactive IgM/IgG in COVID-19 patients. A non-parametric Spearman’s correlation test was used for the statistical analyses. In the graphs, p, r, and N indicate the p-value, correlation coefficient, and sample size, respectively. (C) Distribution of neutralizing activity at different S- and N-specific CLIA-reactive IgM/IgGs. (D) Based on the predicted cutoff value and IgG-S titer, the neutralizing activity of all confirmed patients at different time points was calculated.

To further determine which antibody subclasses and specificities may exert the neutralizing effect, correlations between the titers of the four antibodies and the neutralizing activity were analyzed. The IgG-RBD-S titer demonstrated by far the highest positive correlation with antibody neutralization activity (r=0.6932, p<0.0001), compared to IgM-S (r=0.2220, p<0.05) and IgG-N (r=0.3621, p=0.0001) (Fig. 3B). High levels of neutralizing activity (1:160 or 1:320) were only found in conjunction with high IgG-S, while plasma with high IgG-N titers or unilateral IgM responses did not correlate with high neutralizing activity (Fig. 3C). These findings are consistent with the notion that IgG-S confers neutralizing capacities.

Virus neutralization tests must be performed in BSL3 laboratories which are not broadly available. Therefore, we analyzed the receiver operating characteristic curve and the area under the curve for the IgG-S titers that are associated with virus neutralization. Titers over 4.99 AU/ml were found to constitute a threshold value to predict neutralizing effects, which may help to screen convalescent plasma for immunotherapy if high level biosafety laboratories are not available (Fig. S3). This very strict cut-off value of IgG-S titers was applied to calculate the positive rate of neutralizing activity in samples that could so far not been tested in the neutralization assay. The majority of patients was above this threshold at the latest time point of week 26 indicating the presence of IgG antibodies recognizing the RBD, predictive for neutralizing activity. Please note that our very strict criteria for sensitivity and specificity underestimates the true frequency of individuals with neutralizing antibodies as can be seen when the cut-off is applied to the neutralization data set in figure 3B. Thus, the vast majority of COVID-19 patients raised IgG-RBD-S-binding antibodies with neutralizing capacity, which were maintained over the observational period of 6 months (Fig. 3A & Fig. 3D).

## Discussion

There are tremendous global efforts by companies and academia to design, evaluate, and manufacture prophylactic vaccines against SARS-CoV-2 and the associated COVID-19. Although such a vaccine is obviously highly desirable and first efficacy studies in animals and safety studies in humans appear promising, there is no guarantee that a vaccine will be available soon and that their protection is long-lasting. Another issue is the question, if a natural infection raises a sustained protective immunity, enabling the establishment of collective herd immunity. In both cases, the duration of antibody responses is of critical relevance. At present, the sustainability of protective immunity of convalescent COVID-19 patients is one of the most urgent issues. If existent, convalescent individuals could benefit from their immunity to serve at system relevant positions and long-lasting immunity would also increase the public confidence in vaccines. To the best of our knowledge, with six months, the observational period of our study on the dynamics of antibody responses is the longest so far. We found that SARS-CoV-2-specific IgM recognizing S and N was only transient and disappeared around week 12. Thus, IgM responses will most likely not contribute to sustained immunity against SARS-Cov-2. We even did not find any clear correlation between IgM responses and the ability of plasma to neutralize virus in cell culture. Interestingly, IgG recognizing S and N was maintained at high positive rates and titers for six months. This is particularly important in case of IgG recognizing the RBD of the S protein, which titer correlated with neutralizing activity and was associated with early virus control, highlighting the relevance of IgG-S as correlate of protection in humans. It is important that our patient cohort, which showed sustained IgG responses after a transient contraction phase, exclusively comprised symptomatic COVID-19 patients. The time course as well as the duration of humoral immune responses may well be entirely different following asymptomatic infections ^6^.

Considering that severely ill patients had higher IgG-N levels than non-severe cases at week 4, we speculate that severe COVID-19 patients experience higher virus replication leading to the expression of more virus antigens, which, maybe in combination with a very strong inflammation, elicit strong humoral immune responses persisting for a prolonged period of time. The same hypothesis may explain the transient nature of immunoglobulin responses shown for asymptomatic patients.

It is still controversial how antibody titers and the severity of disease may affect each other. One study found that the total antibody levels in severe patients were significantly higher than those of non-severe patients between the second to fifth week after disease onset, but no differences were observed in IgG or IgM levels alone ^22^. Another study observed that the IgG levels of severe patients were significantly higher than in non-severe patients in the second week after disease onset ^3^. The correlation between high antibody levels and severe COVID-19 brought some discussion if antibodies are involved in immunopathology rather than antiviral effects. Contrary to these studies, we found that in the early period following disease onset in non-severely ill patients and RNA-negative patients (within 3 weeks), the levels of IgM-S/IgM-N/IgG-S/IgG-N were significantly higher than those of severe patients and RNA-positive patients. In addition, there was a clear correlation between IgG-S titers and virus neutralization. This suggests that the antiviral effects of antibodies outweigh potential adverse effects at least during the early phase of COVID-19.

Previous studies have also shown that the plasma of convalescent COVID-19 patients has virus neutralization activity^13^ and alleviates symptoms upon administration to severe patients ^21^. In agreement with previous studies ^12^, we found that IgG-RBD-S is positively correlated with neutralizing activity. However, it was discussed that IgG levels of both symptomatic and asymptomatic patients may decrease rapidly during recovery ^6^, raising concerns about the sustained neutralization activity of patient plasma. Our study demonstrates that the plasma of most symptomatic COVID-19 patients facilitates neutralizing activity during the six months observation period, with a considerable proportion of patients exhibiting very high levels of neutralizing activity.

The discussion of rapidly declining humoral immune responses provoked broad media attention, raising doubts and anxiety about the feasibility of vaccine development and immunity after infection. Based on our data, it appears that the humoral immune response to SARS-CoV-2 in symptomatic COVID-19 patients is rather prototypical for viruses in having an early expansion phase followed by an intermediate contraction phase and a sustained memory phase. Analysis that terminated their observation period earlier than in our study, but extrapolated a long-term trend based on the contraction phase without considering or determining the memory/consolidation phase, bear the inherent risk to come to over-pessimistic conclusions concerning the durability of humoral immune responses after SARS-CoV-2 infection. Even primary infections inducing live-long immunity (e.g. measles infection) and very effective vaccine such as the yellow fever and rabies vaccine have a transient contraction phase in the antibody response. Although only the future will show how long protective immunity will last after natural infections or prophylactic vaccination against SARS-CoV-2, our data suggests that SARS-CoV-2-specific antibody responses are quite similar to responses against many other viruses that induce immunity in humans, including the ‘common-cold’ Corona Viruses that have been shown to mediate protective immunity at for many months to years ^23,24^.

This study has some limitations as follows. First, we did not have enough samples at 9-11 weeks because the patients were placed in mandatory isolation for two more weeks after discharge from the hospital, followed by another two more weeks at home after leaving mandatory isolation. Second, due to the limited availability of the BSL3 laboratory, not all samples could be assessed in virus neutralization tests.

In conclusion, antibodies appear to have antiviral effects in the early stages of SARS-CoV-2 infection; and the most symptomatic patients with COVID-19 remain positive for IgG-S and exhibit sufficient neutralizing activity at six months after the onset of illness. These results support the notion that naturally infected patients have the ability to combat re-infection and vaccines may be able to produce sufficient protection. Please note, that analyses which terminated their observation earlier than ours and extrapolates the long-term trend based on this contraction phase without considering or determining the consolidation phase, bear the inherent risk to come to wrong over-pessimistic conclusions concerning the durability of humoral immune responses.

## Data Availability

All data, models, and code generated or used during the study appear in the submitted article.

## Abbreviations

COVID-19: Corona Virus Disease 2019
SARS-CoV-2: severe acute respiratory syndrome coronavirus 2
SARS: severe acute respiratory syndrome
MERS: middle east respiratory syndrome
ELISA: enzyme-linked Immunosorbent Assay
RBD: receptor-binding domain
RT-PCR: reverse transcription-polymerase chain reaction
PaO2/FiO2: arterial oxygen tension to inspired oxygen fraction
CLIA: capture chemiluminescence immunoassays
VNT: virus neutralization test

## Method

### Patients and sample collection

585 samples obtained from 349 symptomatic COVID-19 patients of the isolation wards or fever clinics of Wuhan Union Hospital or National virus resource Center of Wuhan Institute of Virology, during the period January 1st to July 15th, 2020, were involved in this study. All patients in this study were diagnosed and treated according to the Guidelines of the Diagnosis and Treatment of New Coronavirus Pneumonia (version 7) published by the National Health Committee of the People’s Republic of China^25^. All patients met the following conditions: (1) Epidemiology history, (2) Fever or other respiratory symptoms, (3) Typical CT image abnormities of viral pneumonia, or decreased lymphocyte count, (4) Positive result of IgG and IgM test, or positive result of RT-PCR for SARS-CoV-2 RNA. Severe patients additionally met at least one of the following conditions: (I) low oxygen saturation (≤93%) at resting state, or PaO2 / FiO2≤300mmHg, (2) respiratory failure and requiring mechanical ventilation, (3) multiple organ failure and admittance to an ICU. We retrospectively collected patients’ medical records including demographic factors, laboratory results, and other parameters. Individuals co-infected with human influenza A virus, influenza B virus, or other viruses associated with respiratory infections were excluded. Patients who met at least one of the following conditions: Blood samples were collected and separated by centrifugation at 3000g for 15 min within 4-6 h of collection, followed by 30 min inactivation at 56°C and storage at -20°C for further analyses. This study was approved by the Ethics Commission of Union Hospital of Huazhong University of Science and Technology in Wuhan.

### Detection of SARS-Cov-2 RNA and the IgG and IgM against SARS-CoV-2 S/N

Throat-swab specimens were obtained from all patients and stored in viral-transport medium for SARS-CoV-2 RNA testing. SARS-CoV-2 RNA was detected by real-time reverse transcription polymerase chain reaction (RT-PCR) according to the product manual (Daan gene,Zhongshan, China). Immunoglobulin M (IgM) and G (IgG) antibodies recognizing the SARS-CoV-2 receptor-binding domain (RBD) of the spike (S) or the nucleocapsid (N) protein were tested by capture chemiluminescence immunoassays (CLIA) by MAGLUMI(tm) 2000 Plus (Snibe, Shenzhen, China) as reported ^26^. The cut-off value for IgM-S was 0.7 AU/mL and 1.0 AU/mL for IgG-M, Ig-G, and IgG-S.

### Virus neutralization test (VNT) assay

Vero E6 cells (1×10^4^ per well) were seeded in 96-well plates one night prior to use. Patients’ plasma was incubated at 56°C for 30 minutes to inactivate the complement. Two-fold serially plasma dilutions in the Eagle’s Minimal Essential Medium (EMEM) (NewZongke, Wuhan, China) containing 2% (v/v) fetal bovine serum (FBS) (Gibco, CA, USA) were prepared. SARS-CoV-2 (Strain BetaCoV/Wuhan/WIV04/2019, National Virus Resource Center number: IVCAS 6.7512) at 100 TCID50 was incubated in absence or presence of diluted plasma for 1 h at 37 °C. Afterwards, Vero E6 cell were overlaid with virus suspensions. Each neutralization test was performed in triplicates. At 48 h post infection (p.i.), cytopathic effects (CPE) were visualized and manually judged by microscopic inspection. The neutralizing antibody titer was expressed as the reciprocal value of the highest dilution that prevented CPE formation.

## Statistical Analysis

The mean (standard deviation) was applied for describing continuous variables with a normal distribution, and the median (interquartile range, IQR) was used for continuous variables with a skewed distribution. The difference between groups was examined by Student’s t-test or Mann-Whitney U test, as appropriate. For categorical variables, n (%) was used for description, and examined by Chi-square test or Fisher’s exact test. Dynamic changes of antibodies tracking from day 1 to day 182 after admission were depicted using the locally weighted regression and smoothing scatterplots (Lowess) model (ggplot2 package in R). All statistical analysis was conducted by R (The R Foundation, http://www.r-project.org, version 4.0.0) and SPSS (version 25, IBM, USA). A two-sided P <0.05 was considered statistically significant. The level of statistical significance was depicted as follows: ns, not significance;*p<0.05;**p<0.01;***p<0.001;****p<0.0001.

## Supplementary data

**Figure S1.**
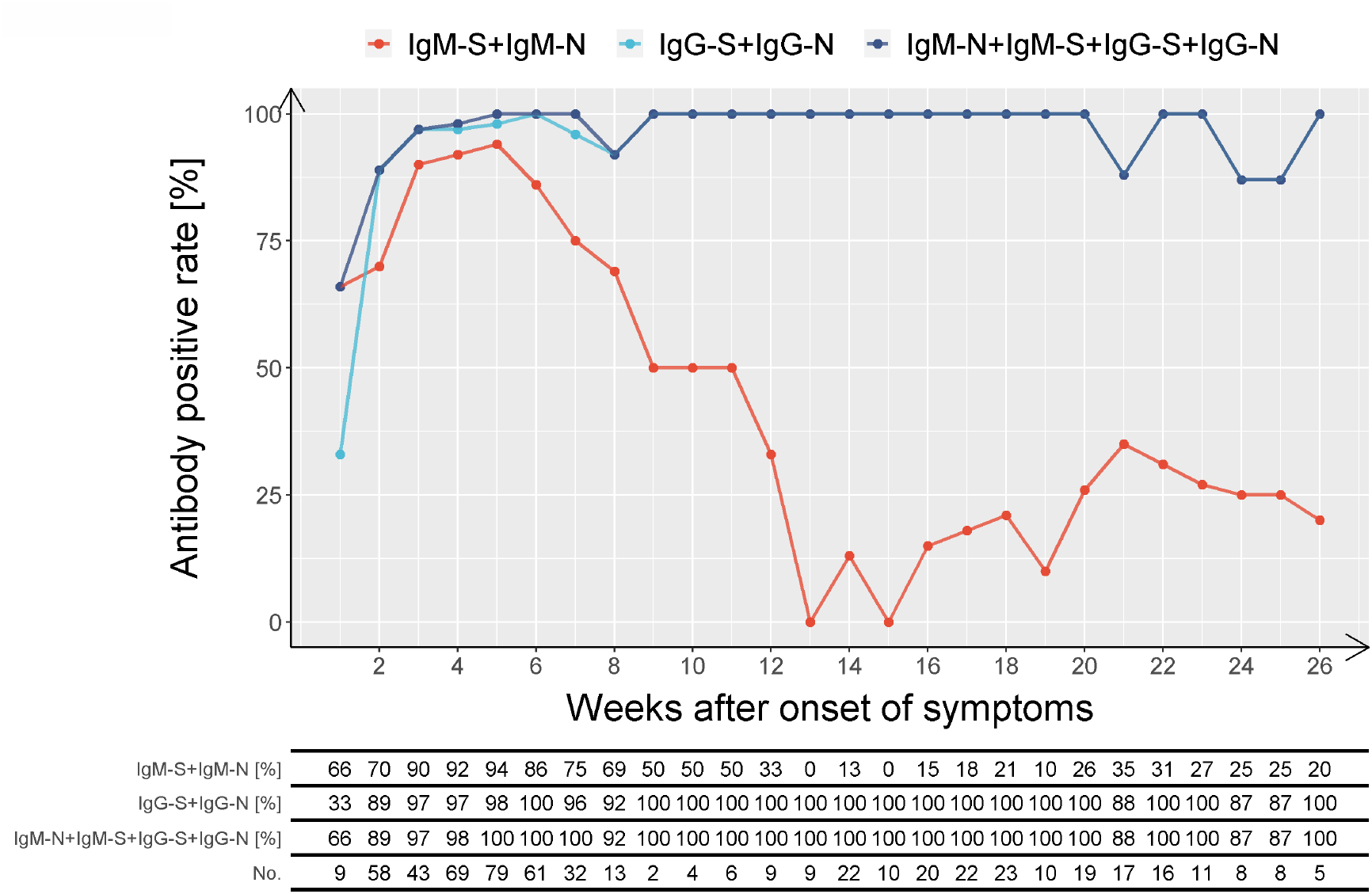
Positive rate of combined antibodies tested in time series following the onset of symptoms. IgM and IgG against the RBD of the spike protein and nucleoproteins of SARS-CoV-2 were detected by capture chemiluminescence immunoassays (CLIA). Positive rate of IgM-S + IgM-N, IgG-S + IgG-N, IgM-S + IgM-N + IgG-S + IgG-N tested in time series after the onset of symptoms.

**Figure S2.**
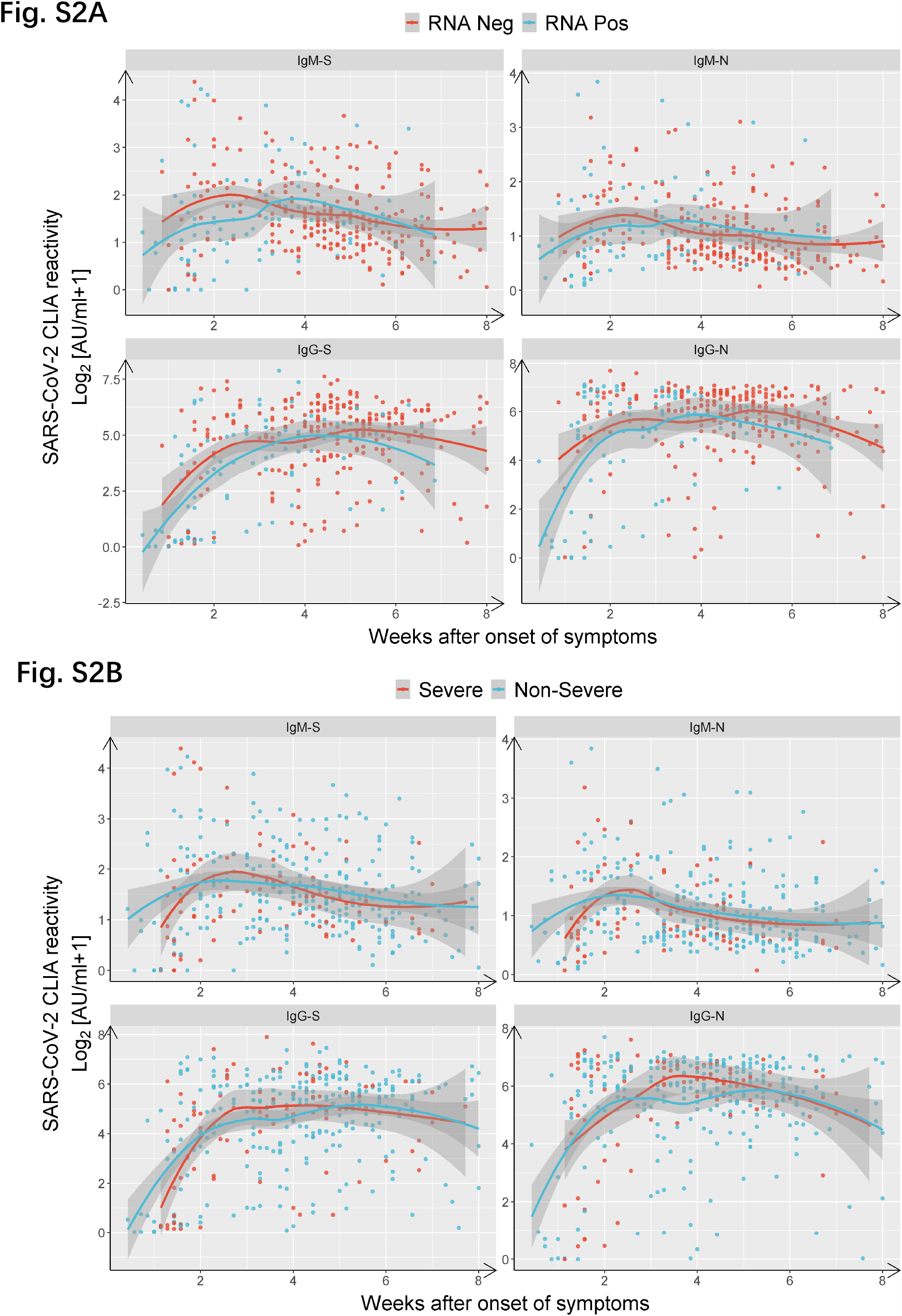
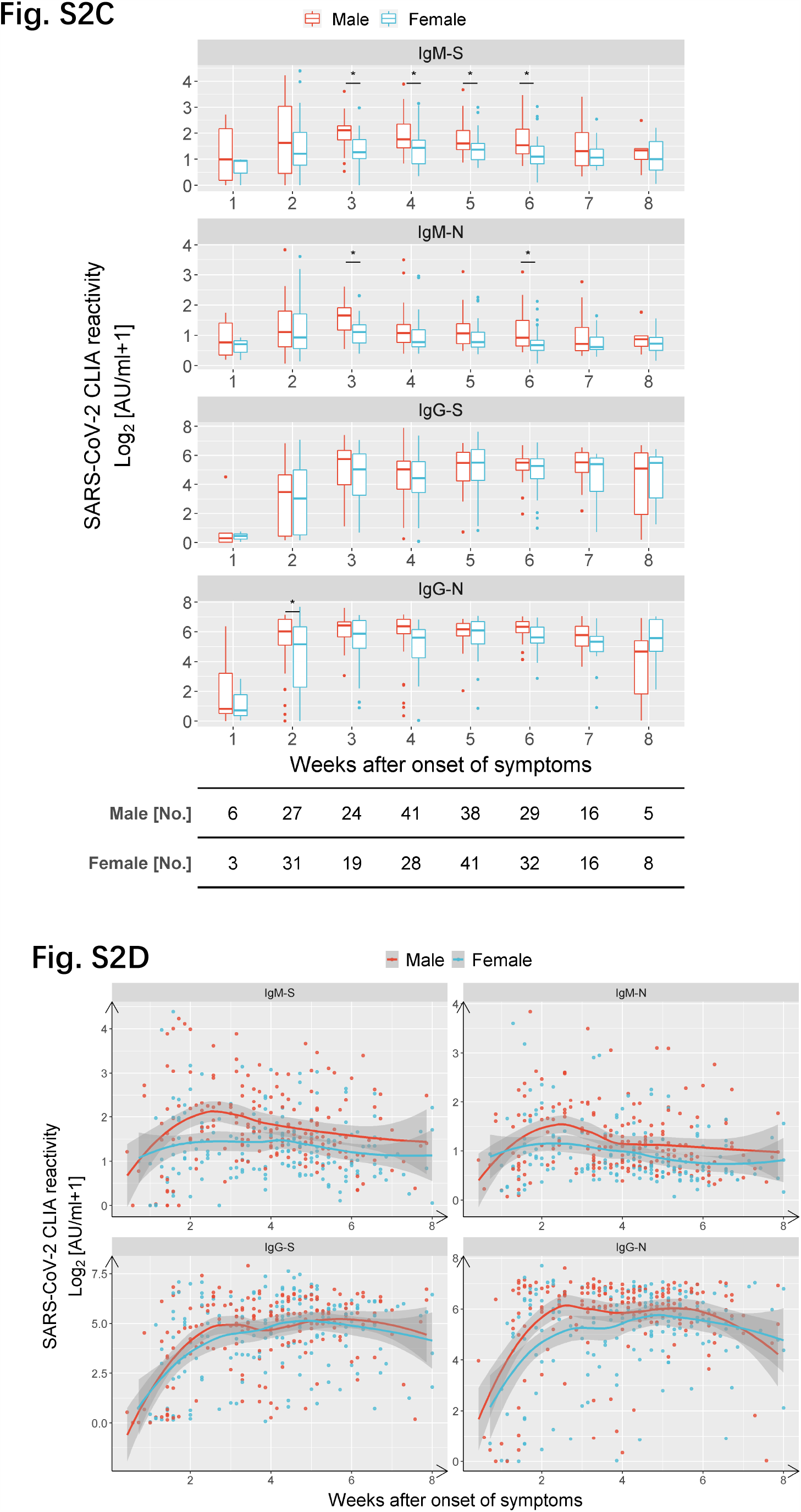
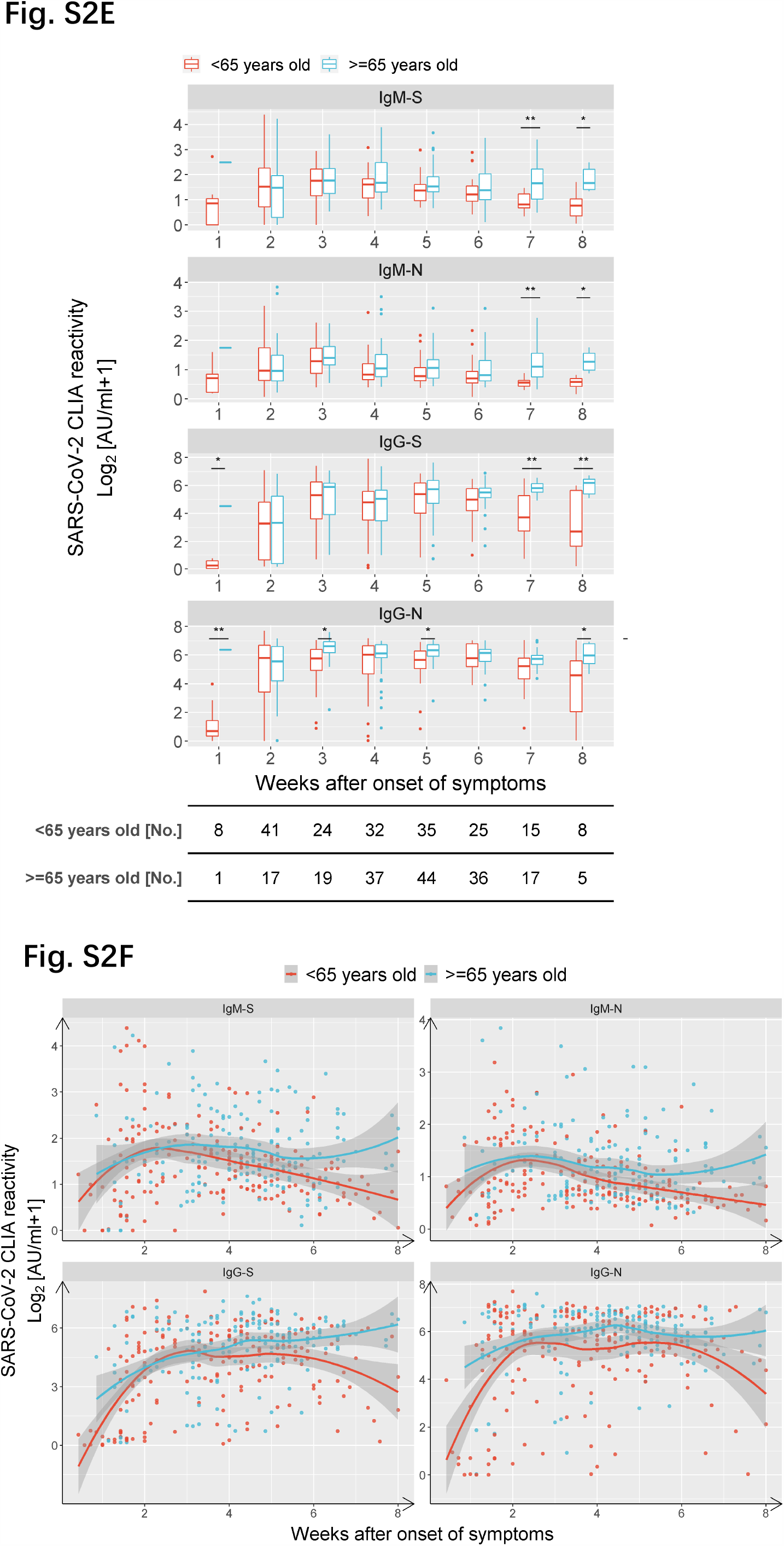
Comparison of differences in antibody titers according to severity of disease, gender, and age by a generalized linear model. Comparison of S and N-specific CLIA-reactive IgM/IgG titers between RNA-positive and -negative cases (A), severe and non-severe patients (B), female and male patients (C-D) and the young (<65 years old) and elder (⩾65 years old) patients (E-F).

**Figure S3.**
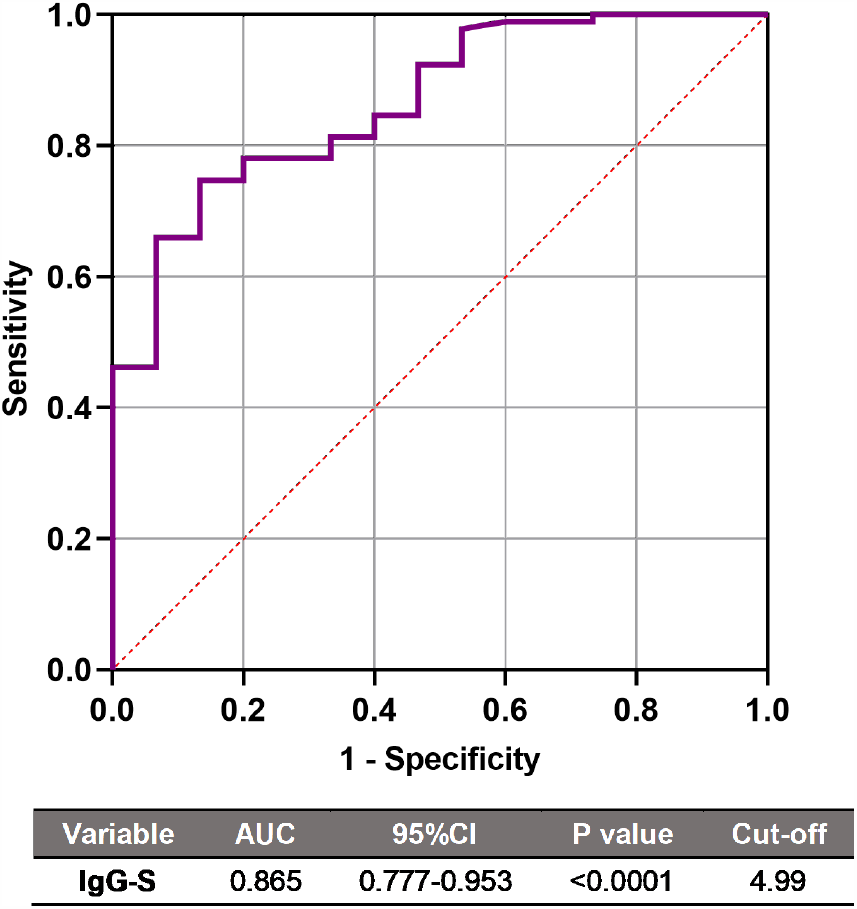
Receiver operating characteristic (ROC) curve of IgG-S titers to predict neutralizing activity of COVID-19 patients. The AUC of IgG-S was 0.865 (95% CI 0.777–0.953; p<0.0001). The optimal cutoff value was 4.99 AU/ml.

**Table S1.** Clinical and laboratory characteristics of hospitalized COVID-19 patients.

| Characteristic | Non-severe, (n=149) | Severe, (n=60) | *P* value |
| --- | --- | --- | --- |
| **Age [years]** | 60.0 (43.5-68.0) | 60.0 (47.3-67.8) | 0.522 |
| **Female, sex [n %]** | 76 (51.0%) | 23 (38.3%) | 0.125 |
| **WBC [$10^9$ /L]** | 5.2 (4.4-6.8) | 5.5 (4.2-7.9) | 0.199 |
| **N [$10^9$ /L]** | 3.3 (2.7-4.5) | 4.4 (2.8-6.3) | **0.004** |
| **L [$10^9$ /L]** | 1.4 (1.0-1.7) | 0.9 (0.6-1.2) | **0.000** |
| **M [$10^9$ /L]** | 0.5 (0.4-0.7) | 0.5 (0.3-0.6) | 0.546 |
| **N [%]** | 63.3 (55.4-70.9) | 71.7 (59.1-79.9) | **0.001** |
| **L [%]** | 25.3 (19.2-33.7) | 20.1 (11.8-31.1) | **0.005** |
| **M [%]** | 8.9 (6.9-11.0) | 7.3 (5.2-9.3) | **0.001** |
| **PLT [$10^9$ /L]** | 211.0 (170.5-275.0) | 200.5 (143.8-236.8) | **0.029** |
| **TBil [μmol /L]** | 10.9 (9.0-14.3) | 12.9 (9.7-16.5) | **0.021** |
| **ALT [U/L]** | 25.0 (18.0-45.0) | 37.0 (24.0-54.0) | **0.007** |
| **AST [U/L]** | 24.0 (19.0-34.5) | 40.0 (28.0-56.0) | **0.000** |
| **LDH [U/L]** | 224.0 (181.0-288.0) | 355.0 (271.5-449.0) | **0.000** |
| **CK [U/L]** | 62.0 (43.0-88.0) | 76.0 (48.5-200.5) | **0.016** |
| **Cr [μmol /L]** | 67.4 (55.7-79.0) | 72.0 (58.0-86.4) | **0.046** |
| **D-Dimer [mg/L]** | 0.5 (0.2-1.0) | 0.9 (0.4-2.9) | **0.001** |
| **PT [s]** | 13.2 (12.7-13.8) | 13.6 (12.9-14.4) | **0.030** |
| **TT [s]** | 17.6 (16.6-18.5) | 17.5 (16.6-18.6) | 0.785 |
| **APTT [s]** | 37.0 (35.1-40.0) | 37.7 (35.0-41.4) | 0.436 |
| **FIB [g/L]** | 4.2 (3.4-5.1) | 5.4 (4.4-6.7) | **0.000** |

PLT: platelet, WBC: white blood cells, N: neutrophil count, M: monocyte count, L: lymphocyte count, N%: neutrophil percentage, M%: monocyte percentage, L%: lymphocyte percentage, total bilirubin (TBil), ALT: alanine aminotransferase, AST: aspartate aminotransferase, CK: creatine kinase, LDH: lactate dehydrogenase, Cr: Creatinine and prothrombin time, PT.
All data are presented as the median (IQR) or n (%).
All data are calculated applying a Mann-Whitney U test or $\chi^2$ test.
P <0.05 was considered statistically significant.

